# Trends in Pediatric Firearm-Related Encounters during the COVID-19 Pandemic by Age Group, Race/Ethnicity, and Schooling Mode in Tennessee

**DOI:** 10.1101/2023.03.17.23287413

**Authors:** Tara McKay, Kelsey Gastineau, Jesse O. Wrenn, Jin H. Han, Alan B. Storrow

## Abstract

**Purpose:** Increases in pediatric firearm-related injuries during the COVID-19 pandemic may be due to changes in where children and adolescents spent their time. This paper examines changes in the frequency of pediatric firearm-related encounters as a function of schooling mode overall and by race/ethnicity and age group at a large trauma center through 2021.

**Methods:** We use data from a large pediatric and adult trauma center in Tennessee from January 2018 to December 2021 (N=211 encounters) and geographically linked schooling mode data. We use Poisson regressions to estimate smoothed monthly pediatric firearm-related encounters as a function of schooling mode overall and stratified by race and age.

**Results:** Compared to pre-pandemic, we find a 42% increase in pediatric encounters per month during March 2020 to August 2020, when schools were closed, and a 23% increase in encounters after schools returned to in-person instruction. Effects of schooling mode are heterogeneous by race. Encounters increased among non-Hispanic Black children and adolescents across all periods relative to pre-pandemic. Among non-Hispanic white children and adolescents, encounters increased during the closure period and decreased on return to in-person instruction. Effects of schooling mode are also heterogeneous by age. Relative to pre-pandemic, pediatric firearm-related encounters increased 205% for children aged 5 to 11 and 69% for adolescents aged 12 to 15 during the school closure period.

**Conclusion:** COVID-19-related changes to school instruction mode in 2020 and 2021 are associated with changes in the frequency and composition of pediatric firearm-related encounters at a major trauma center in Tennessee.

## Introduction

From 2019 to 2020, firearm-related deaths among children and adolescents increased by almost 30%, making firearms the leading cause of death among young people in the United States.^1^ Although this increase may be partially driven by prevailing trends,^2^ other evidence suggests that some of this increase is related to the COVID-19 pandemic.^3–6^

The reasons for this increase remain underexamined empirically. Recent work proposes several potential drivers, including changes in where children spent their time; decreased parental supervision; increased firearm purchasing, neighborhood violence, and child abuse; and reluctance among adolescents to socialize outside the home.^7,8^ This study examines whether changes in schooling mode (e.g, in-person, virtual, hybrid, closed) are associated with changes in the frequency of pediatric firearm-related encounters overall and by race/ethnicity and age group at a large trauma center in Tennessee.

Schooling mode provides a proxy measure of where and how children and adolescents spent their time during the pandemic. Changes in schooling mode may have increased risk of pediatric firearm injury by increasing time spent at home. About 40% of U.S. children live in homes with at least one firearm. In Tennessee, nearly half (46% as of 2016) of households own a gun.^9^ In over half of American gun-owning households, firearms are not stored locked and unloaded.^10^ Firearm storage practices strongly predict risk of firearm homicide, suicide, and unintentional injury among children and adolescents.^11^

The effects of changes in schooling mode on firearm injury may vary by race/ethnicity. While firearms are present for youth from all racial/ethnic backgrounds, white households with children are more likely to report owning a firearm compared with Black households.^12,13^ Firearm injury among white children and adolescents is more likely to occur in the home and be self-inflicted^14,15^ or perpetrated by a parent/caregiver^16^ compared with Black children, and is predicted by household firearm ownership.^2,12^ However, both handgun ownership in Black households and firearm suicide among Black male adolescents have increased in recent years.^13^ Some research finds differences in household firearm storage practices by race, with white households reporting less safe storage practices than Black households.^17^ School closures and virtual/hybrid schooling modes may have therefore differentially increased risk of exposure to firearms for white children and adolescents compared to Black children and adolescents.

Conversely, in 2020, Black adolescents were nearly 10 times more likely to experience firearm homicide than their white peers.^18^ These disparities are the result of structural racism, which increases the likelihood that Black adolescents live in neighborhoods with concentrated poverty, unemployment, residential instability, and gun violence.^19–21^ Black children and adolescents have higher rates of firearm injury relative to the population that are increasing faster than other racial/ethnic groups.^2,15^ In the context of the COVID-19 pandemic, risk of firearm injury among Black children and adolescents may be less directly affected by schooling mode changes because Black children and adolescents are exposed to firearms both in and out of the home.

Increased time at home may also have differential effects on firearm injury by age. For younger children aged 0 to 12, 85% of firearm deaths occur in the home.^14^ While a majority of older children and adolescents who die by firearm are also shot in the home, fatal firearm homicides among adolescents aged 13 to 17 years are equally likely occur at home (39%) versus on the street/sidewalk (38%).^14^ Pandemic-related changes to schooling that increased time at home may have increased accidental and unintentional firearm injuries among all ages, especially younger children. There may also have been changes in the frequency of firearm suicide among adolescents; however, research suggests that increased access to a firearm at home and worsening mental health may have increased firearm suicide^22,23^ while disruption of bullying and academic stressors decreased firearm suicide among this age group.^24^

We advance current literature by examining the effects of schooling mode on the frequency of pediatric firearm-related encounters overall and by age and race/ethnicity using data from a large pediatric trauma center in Tennessee. We hypothesize that school closures differentially affected pediatric firearm encounters by race/ethnicity, with greater increases for white children, and by age, with greater increases for younger children.

## Methods

To examine changes in the frequency of pediatric firearm-related encounters, we used data on Hospital and Emergency Department firearm-related encounters at Monroe Carell Jr. Children’s Hospital at Vanderbilt University (MCJCHV) and Vanderbilt University Medical Center (VUMC). MCJCHV and VUMC are one of a small number of pediatric and adult Level I Trauma Centers in Tennessee and the only pediatric and adult Level I Trauma Centers in the Middle Tennessee region, one of the three administrative regions of Tennessee encompassing 33 counties and the most populous city, Nashville. MCJCHV and VUMC maintain a service area of 80,000 square miles, which is home to 1.25 million children in 79 counties across Tennessee, southern Kentucky, and northern Alabama.

Trauma patients under age 16 are triaged to the Pediatric Emergency Department at MCJCHV. Trauma patients aged 16 or 17 years old who have life-threatening injuries may be triaged to the Adult Emergency Department at VUMC. This study was reviewed by the Institutional Review Board at VUMC.

### Study Cohort

We queried the institutional database of patient encounters for initial firearm-related Emergency Department encounters at both hospitals from January 1, 2018, through December 31, 2021. Because firearm-related diagnosis codes are used inconsistently, encounters were initially identified via text search of “gun,” “GSW,” “firearm” and variations in the Epic diagnosis field. Codes and diagnoses that suggested historical firearm-related injuries were excluded (e.g., History of Gunshot Wound, Healing Gunshot Wound). All diagnoses were manually reviewed by two members of the study team and diagnoses unrelated to firearm injuries were removed. This yielded 211 pediatric patients aged 5-17 years.

### Measures

The primary outcome was the number of monthly pediatric firearm-related encounters overall, by race/ethnicity, and by age group. Monthly values were smoothed using 3-month moving average.

For each encounter, we also captured information on patient age, race, ethnicity, sex, diagnosis, and billing ZIP code. Race was recorded as white, Black or African American, other, unknown/missing, or refused. Ethnicity was recorded as “Hispanic, Latino/a, or Spanish,” “Not Hispanic, Latino/a, or Spanish,” unknown/missing, or refused. We coded race/ethnicity as “non-Hispanic Black,” “non-Hispanic white,” and “other” due to the small number of encounters among Hispanic/Latino children and adolescents (N=8). Encounters with missing data on race/ethnicity are included in overall and age stratified analyses.

We established the primary schooling mode for each month using MCH Strategic Data^25^ for Fall 2020, Spring 2021, and Fall 2021 linked via ZIP-to-School-District crosswalk to our institutional encounter data. MCH schooling mode data have been used in studies of COVID-19 in K-12 schools in the US.^26^ In the US, most K-12 schools closed by the weeks of March 16 or March 23, 2020, and remained closed or provided online schooling for the remainder of the school year.^27^ During Summer 2020, about 60% of summer camps and community programs for youth were closed across the United States.^28^ In Fall 2020, school districts adopted a variety of schooling modes, with about half offering hybrid instruction, 24% online only, and 17% in-person only.^25^ Many districts also delayed the start of the 2020-2021 academic year by 1 to 6 weeks. Schooling mode remained varied in Spring 2021, with some of the country’s largest districts remaining virtual for most or all of Spring 2021 while others reinstated in-person instruction between January and March.

In Tennessee, the timing of schooling mode changes for the largest public-school districts is highly consistent with national trends. In Fall 2020, most districts adopted a hybrid or mixed mode of instruction. A minority offered in-person only instruction or online only instruction. Several districts delayed the start of the academic year, began the year fully online, or closed due to COVID-19 cases in Fall 2020. In Spring 2021, most districts returned to in-person instruction by the end of the term.

Three-quarters (73%) of patients presenting to MCJCHV or VUMC for pediatric firearm-related encounter from 2018 to 2021 lived in 4 of Tennessee’s 95 counties: Nashville-Davidson (53.7%), Rutherford (8.8%), Montgomery (5.6%), and Sumner (5.1%). Review of schooling mode for the school districts serving these 4 counties established the following timings: pre-pandemic, 01/2018 – 02/2020; closed, 03/2020 – 08/2020; virtual/hybrid, 09/2020 – 02/2021; in-person, 03/2021 – 12/2021.

### Analyses

All analyses were restricted to pediatric patients aged 5 to 17 years. We began with descriptive statistics and time trends. We then used a series of Poisson regressions with month-of-year fixed effects and robust standard errors to predict smoothed monthly pediatric firearm-related encounters as a function of schooling mode overall and stratified by race/ethnicity and age.

We tested for overdispersion in all models and consistently found that Poisson models are appropriate. Findings are robust to +/- 1 month shifts in the timing of school mode changes, the exclusion of March 2020, and the censoring of four encounters from a school shooting in Kentucky in January 2018 shown on Figure 1 (see Appendix A).

**Figure 1.**
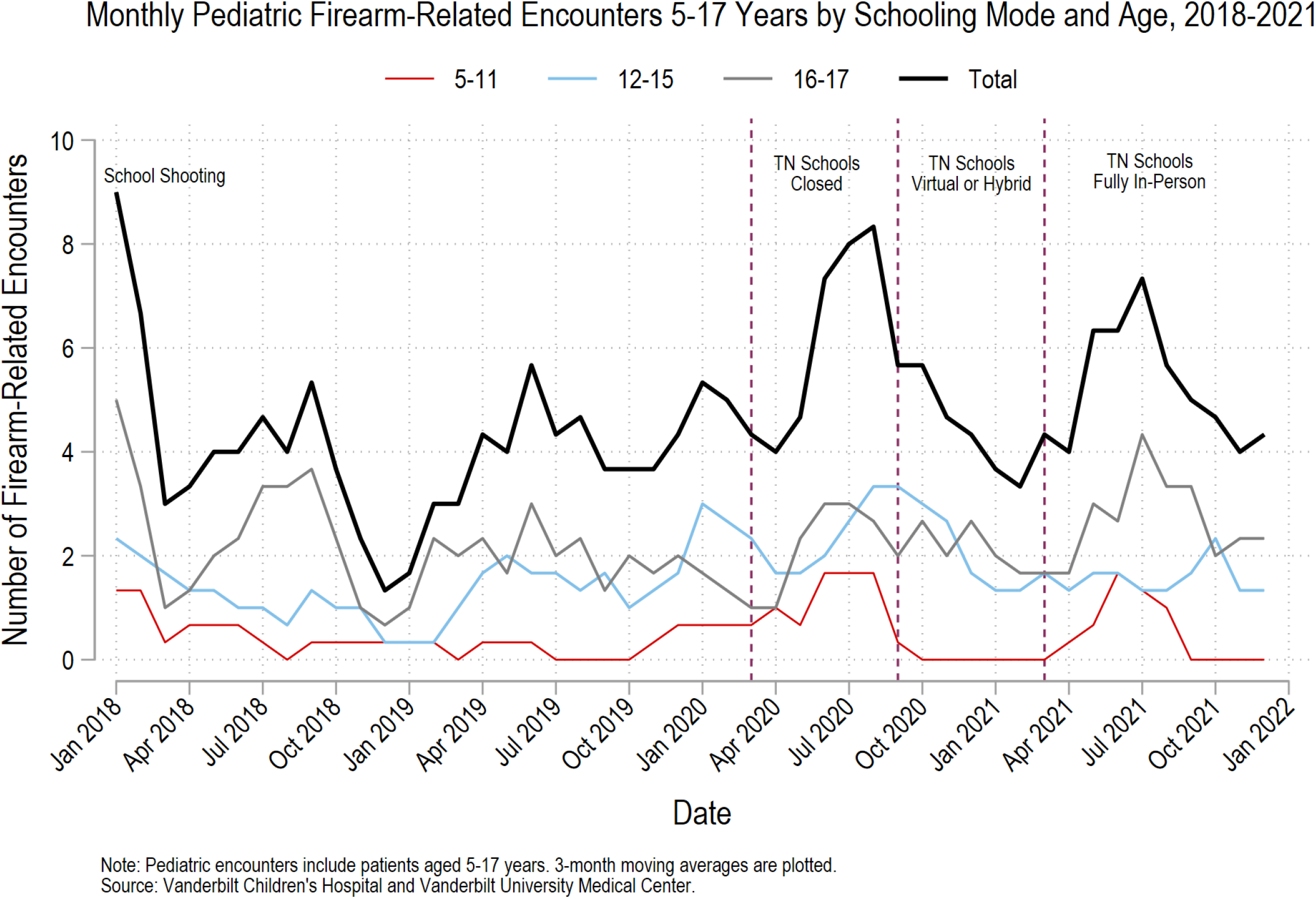
Monthly Pediatric Firearm-related Encounters by Schooling Mode and Age, 2018-2021

### Patient and Public Involvement

No patients were involved in the design or conduct of the research.

## Results

Table 1 presents patient characteristics for pediatric firearm-related encounters from January 2018 through December 2021. Consistent with national trends, most pediatric patients presenting for a firearm-related encounter were male across all time periods. Adolescents aged 16 and 17 comprised a majority of encounters in all periods except the school closure period (March to August 2020). Non-Hispanic Black pediatric patients comprised an increasing share of encounters, growing from 41% in the pre-pandemic period to 62% following the return to in-person instruction in March 2021.

**Table 1.**
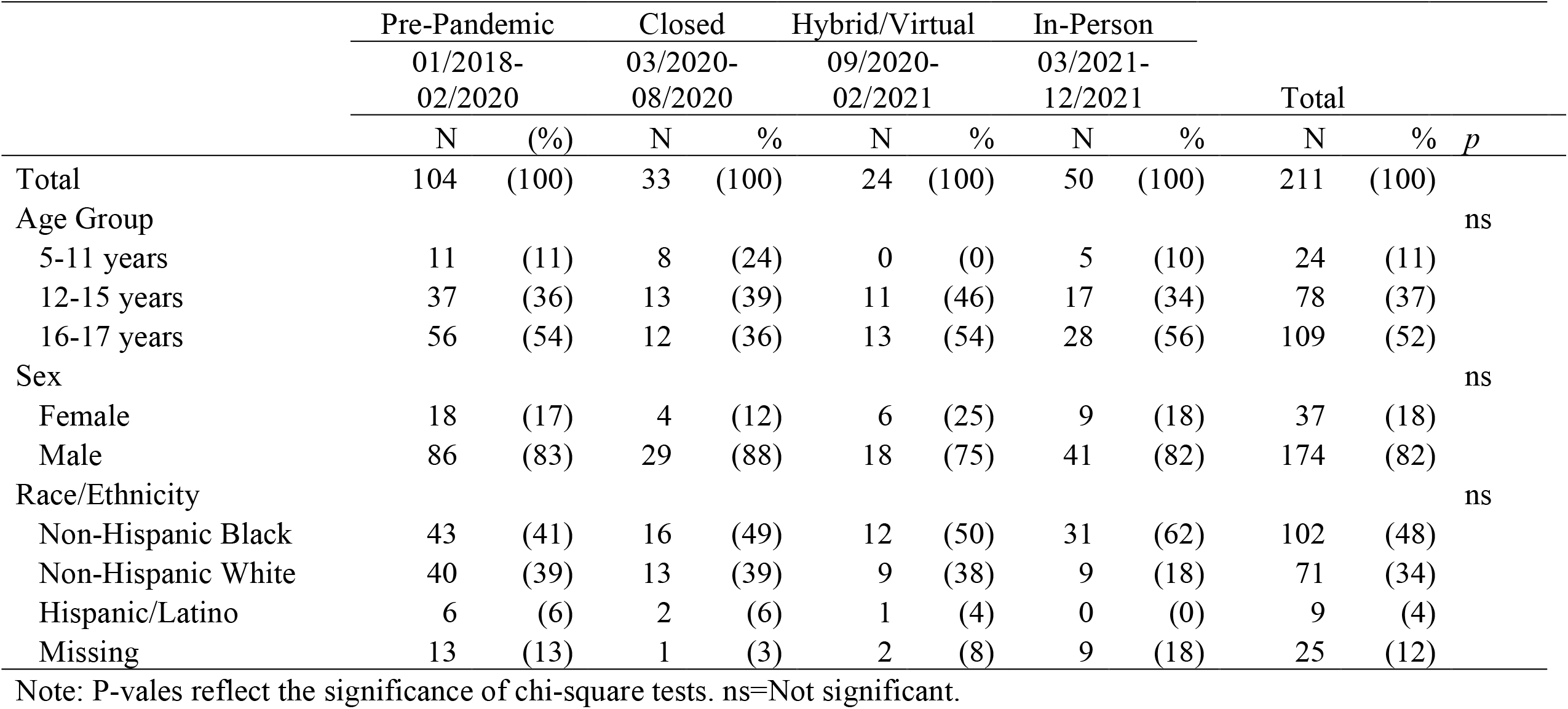
Demographic and Encounter Characteristics for Pediatric Firearm-Related Encounters, by Schooling Mode Period

|  | Pre-Pandemic |  | Closed |  | Hybrid/Virtual |  | In-Person |  |  |  |  |
| --- | --- | --- | --- | --- | --- | --- | --- | --- | --- | --- | --- |
|  | 01/2018-02/2020 |  | 03/2020-08/2020 |  | 09/2020-02/2021 |  | 03/2021-12/2021 |  | Total |  |  |
|  | N | (%) | N | % | N | % | N | % | N | % | *p* |
| Total | 104 | (100) | 33 | (100) | 24 | (100) | 50 | (100) | 211 | (100) |  |
| Age Group |  |  |  |  |  |  |  |  |  |  | ns |
| 5-11 years | 11 | (11) | 8 | (24) | 0 | (0) | 5 | (10) | 24 | (11) |  |
| 12-15 years | 37 | (36) | 13 | (39) | 11 | (46) | 17 | (34) | 78 | (37) |  |
| 16-17 years | 56 | (54) | 12 | (36) | 13 | (54) | 28 | (56) | 109 | (52) |  |
| Sex |  |  |  |  |  |  |  |  |  |  | ns |
| Female | 18 | (17) | 4 | (12) | 6 | (25) | 9 | (18) | 37 | (18) |  |
| Male | 86 | (83) | 29 | (88) | 18 | (75) | 41 | (82) | 174 | (82) |  |
| Race/Ethnicity |  |  |  |  |  |  |  |  |  |  | ns |
| Non-Hispanic Black | 43 | (41) | 16 | (49) | 12 | (50) | 31 | (62) | 102 | (48) |  |
| Non-Hispanic White | 40 | (39) | 13 | (39) | 9 | (38) | 9 | (18) | 71 | (34) |  |
| Hispanic/Latino | 6 | (6) | 2 | (6) | 1 | (4) | 0 | (0) | 9 | (4) |  |
| Missing | 13 | (13) | 1 | (3) | 2 | (8) | 9 | (18) | 25 | (12) |  |

Note: P-values reflect the significance of chi-square tests. ns=Not significant.

Figure 1 presents the 3-month moving average of monthly pediatric gunshot wound encounters from January 1, 2018, to December 31, 2021. Vertical lines indicate pandemic-related changes in the mode of instruction. In the pre-pandemic period, the trauma center saw an average of 3.9 pediatric encounters among 5- to 17-year-olds monthly. The trauma center experienced an increase in pediatric firearm-related encounters during the school closure period in 2020 to an average of 5.6 encounters per month by late summer 2020. The 2020 peak is followed by a decline in encounters to 4.5 per month on average during the hybrid/virtual period, followed by a rise in encounters to 4.8 per month on average after return to in-person instruction in early Spring 2021.

Table 2 presents the predicted number of monthly pediatric firearm-related encounters and percent change from pre-pandemic period controlling for month-of-year fixed effects overall and stratified by race/ethnicity and by age. Compared to the pre-pandemic period (January 2018 to February 2020), the trauma center saw a significant increase of 1.7 pediatric firearm-related encounters per month, a 42% increase over baseline (*p*<0.001), from March 2020 to August 2020 when schools and other youth activities were closed or canceled. During the virtual/hybrid schooling period, the number pediatric firearm-related encounters remained elevated relative to baseline, increasing by 15% over pre-pandemic (*p*<0.1). During the in-person schooling period beginning in March 2021, pediatric firearm-related encounters increased by 0.9 encounters per month over baseline to 4.8 encounters on average, or 23% (*p*<0.001).

**Table 2.** Predicted Number of Monthly Pediatric Firearm-Related Encounters and Percent Change Relative to Pre-Pandemic from Adjusted Poisson Models, Overall and Stratified by Race/Ethnicity and Age Group

| School Mode | By Race/Ethnicity |  |  |  |  |  |  |  |  |
| --- | --- | --- | --- | --- | --- | --- | --- | --- | --- |
|  | Overall<sup>a</sup> |  |  | Non-Hispanic Black |  |  | Non-Hispanic White |  |  |
|  | Number<br>per<br>Month | % | *p* | Number<br>per<br>Month | % | *p* | Number<br>per<br>Month | % | *p* |
|  |  | Change<br>from Pre-Pandemic |  |  | Change<br>from Pre-Pandemic |  |  | Change<br>from Pre-Pandemic |  |
| Pre-Pandemic (1/2018-2/2020) | 3.9 | -- |  | 1.6 | -- |  | 1.5 | -- |  |
| Closed (3/2020-8/2020) | 5.6 | 42 | *** | 2.8 | 70 | *** | 1.9 | 29 | * |
| Virtual/Hybrid (9/2020-2/2021) | 4.5 | 15 |  | 2.4 | 48 | * | 1.8 | 19 |  |
| In-Person (3/2021-12/2021) | 4.8 | 23 | *** | 3.0 | 80 | *** | 0.8 | -45 | *** |

| School Mode | By Age Group |  |  |  |  |  |  |  |  |
| --- | --- | --- | --- | --- | --- | --- | --- | --- | --- |
|  | Age 5-11 |  |  | Age 12-15 |  |  | Age 16-17 |  |  |
|  | Number<br>per<br>Month | % | *p* | Number<br>per<br>Month | % | *p* | Number<br>per<br>Month | % | *p* |
|  |  | Change<br>from Pre-Pandemic |  |  | Change<br>from Pre-Pandemic |  |  | Change<br>from Pre-Pandemic |  |
| Pre-Pandemic (1/2018-2/2020) | 0.4 | -- |  | 1.4 | -- |  | 2.1 | -- |  |
| Closed (3/2020-8/2020) | 1.2 | 205 | *** | 2.4 | 69 | *** | 2.1 | -2 |  |
| Virtual/Hybrid (9/2020-2/2021) | 0.1 | -85 |  | 2.2 | 55 | * | 2.2 | 5 |  |
| In-Person (3/2021-12/2021) | 0.6 | 61 |  | 1.6 | 14 |  | 2.7 | 26 | ** |

Note: \* $p < 0.05$ , \*\* $p < 0.01$ , \*\*\* $p < .001$ . <sup>a</sup> All models are predicted using a Poisson model with month of year fixed effects and robust standard errors.

We observe differences in the direction and magnitude of schooling mode effects by race/ethnicity and age (see Figure 2). Regression models stratified by race show that firearm-related encounters increased significantly relative to pre-pandemic among non-Hispanic Black patients in all periods by a substantial margin: 70% in the school closure period, 48% in the hybrid/virtual period, and 80% after the return to in-person instruction. For non-Hispanic white children and adolescents, firearm-related encounters increased 29% during the school closure period and then decreased 45% after the return to in-person instruction relative to pre-pandemic. Compared to all other groups, non-Hispanic white children and adolescents were significantly less likely to present for a firearm-related encounter after schools returned to in-person instruction (*p*<.05).

**Figure 2.**
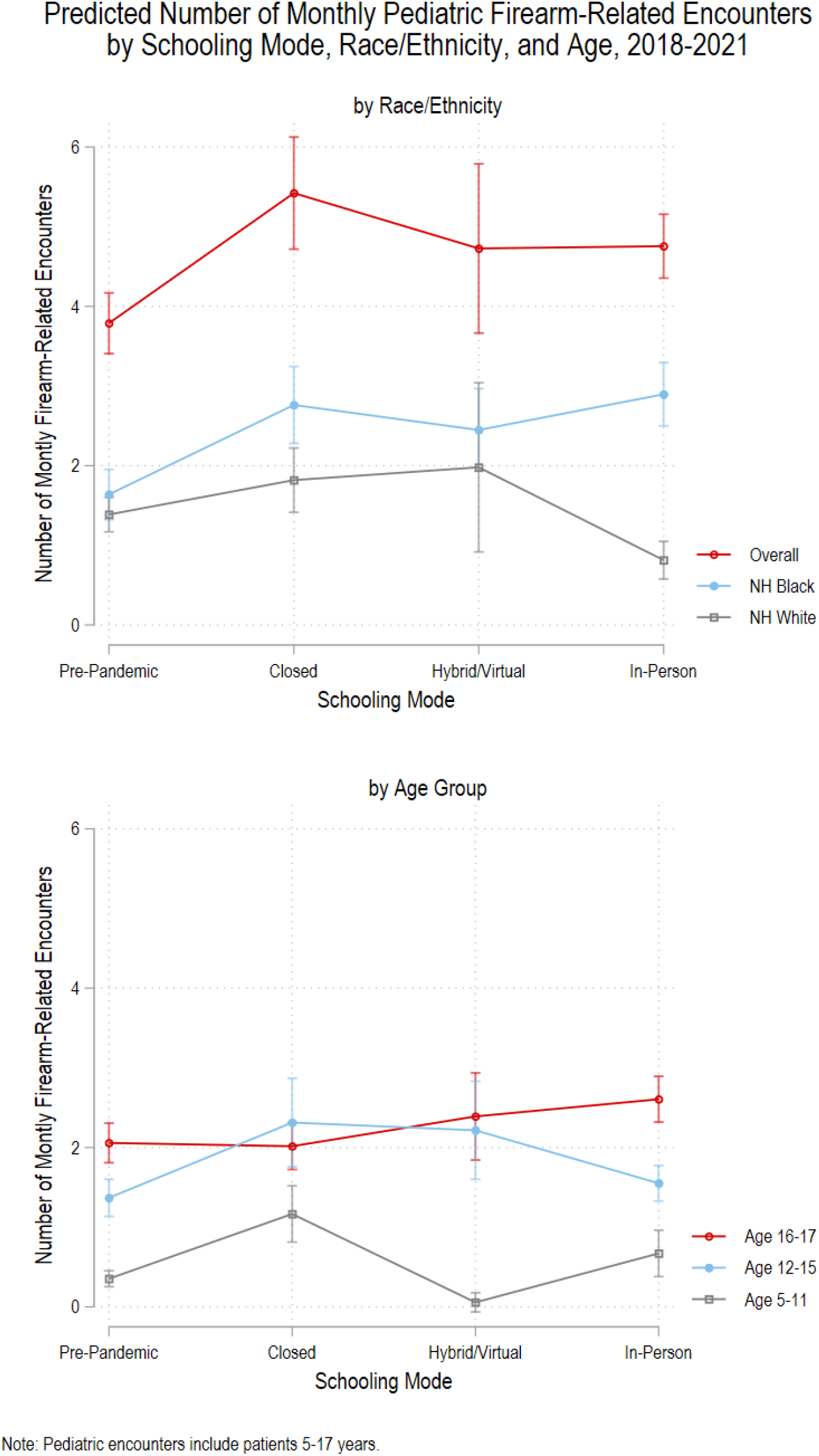
Predicted Number of Monthly Pediatric Firearm-Related Encounters by Schooling Mode, Race/Ethnicity, and Age, 2018-2021

Regression models stratified by age show that pediatric firearm-related encounters increased significantly among children aged 5 to 11 and 12 to 15 during the school closure period. Relative to pre-pandemic, pediatric firearm-related encounters increased by 205% for children aged 5 to 11 and 69% for adolescents aged 12 to 15 during the school closure period. The rate of encounters remained elevated during the virtual/hybrid period for adolescents 12 to 15 years old. Firearm encounters did not change relative to baseline among adolescents aged 16 and 17 until after the return to in-person instruction, where we observe a 26% increase in firearm encounters.

## Discussion

We examine trends in pediatric firearm-related encounters at a single center from January 1, 2018, to December 31, 2021. Our analyses extend existing work by 1) linking information on schooling mode to examine associations with changes in the number of pediatric firearm-related encounters; 2) including the second full academic year of the pandemic; and 3) stratifying by race/ethnicity and age to assess diverging trajectories of firearm risk.

Trends in pediatric firearm-related encounters were closely tied to changes in schooling mode. The 42% increase in monthly pediatric firearm-related encounters during March 2020 to August 2020, when schools were closed and other youth activities canceled, is highly consistent with the 38% increase in firearm-related encounters reported using data from the same period for 44 US children’s hospitals in the Pediatric Health Information System database.^5^ In Fall 2020, when most schools in the service area provided hybrid or virtual schooling, the rate of encounters was not statistically different from baseline. As students returned to in-person instruction in early Spring 2020, monthly pediatric firearm-related encounters again increased over the pre-pandemic period.

Importantly, however, the 23% increase observed during the return to in-person period obscures a divergence in the frequency of firearm injury by race/ethnicity. Although white and Black children experienced more firearm injuries during the school closure period relative to pre-pandemic, firearm injuries decreased well below baseline (−45%) following the return to in-person instruction for white children while injuries continued to escalate for Black children. Although prior work found no significant changes in the racial/ethnic distribution of pediatric firearm-related encounters from March 2020 to August 2020,^5^ which our findings also support, we provide new evidence of diverging trajectories by race/ethnicity as the pandemic continued.

These findings are consistent with other work documenting greater and increasing risks of firearm injury death among Black children and adolescents who experience a broader set of household and community gun violence risks.^2,29,30^ While this study does not examine underlying causes, evidence has shown that socioeconomically disadvantaged neighborhoods, a result of persistent structural and systemic racism, are associated with larger Black-white disparities in firearm injuries and fatalities.^21^ Many root causes of gun violence including income inequality, poverty, and funding of public housing were exacerbated by the COVID-19 pandemic.^20,31^

We also find evidence of changes in the frequency of pediatric firearm-related encounters by age. The monthly number of encounters among children aged 5 to 11 and 12 to 15 increased significantly during the school closure period compared to pre-pandemic. These findings are consistent with expectations that children and younger adolescents spent more time at home where a firearm may have been present during the pandemic. Firearm injuries and deaths in younger children are more likely to be unintentional shootings^2,14^ and involve a firearm originating from either from the child’s home or the home of a friend or relative.^32^

As schools returned to in-person instruction, encounters among 16- and 17-year-olds increased by 26%. This is consistent with other work showing a 12 to 18% increase in teen suicides following the return to in-person instruction in Spring 2021.^24^ The apparent protectiveness of school closures and virtual/hybrid instruction for 16- and 17-year-olds may reflect the accumulation of mental health challenges wrought by the pandemic that only became apparent over time, more frequent contact with family members, or the interruption of bullying and academic stressors by school closures and hybrid/virtual instruction.^23,24^

For children and adolescents living in a house with a firearm, storage practices matter. Research consistently finds that storing a firearm locked and unloaded and storing ammunition separately are independently effective practices for reducing self-inflicted and unintentional firearm injuries among children and adolescents.^14,32^

These findings highlight the pandemic as a critical opportunity to improve firearm safe storage counseling. Despite healthcare providers’ understanding of the public health impact of firearm injuries and importance of counseling, many do not regularly or consistently ask about firearm ownership and storage in patient encounters.^33^ Many barriers, including lack of time, uncertainty of the effect, inadequate training, and perceived parental resentment, have been cited for decades. ^34,35^ However, most parents are generally open to these conversations,^36^ and physician counseling has been shown to be effective in producing safer storage practices at home, especially when a secure storage device such as a cable gun lock is provided.^37–39^ The American Academy of Pediatrics and others have recommended counseling about firearm safety and gun violence for over a decade.^40,41^

This study has limitations. As a single center study, the total number of pediatric firearm-related encounters observed is small (N=211), limiting generalizability to other areas or institutions. However, our findings are consistent with others documenting substantial increases in firearm injuries and deaths during the pandemic.^4–6^ Additionally, we use the timing of schooling mode changes that applies to a majority of encounters, which may obscure finer variation for a minority originating from districts that returned to in-person instruction slightly earlier (mid-Fall 2020) or later (late-Spring or Fall 2021). Misclassification of race/ethnicity is unlikely to be random and may attenuate effects for non-Hispanic Black children and contribute to few encounters observed among other minority groups. Last, we used schooling mode as a proxy for where and how children and adolescents spent their time. We are unable to observe how these changes affected parental supervision, intent, or other proposed determinants of increased firearm-related injury and death during the pandemic (e.g., increased child abuse). Further, other mechanisms, including increased purchasing of firearms during the pandemic^3^ and in response to other events (e.g., Black Lives Matter protests and the 2020 National Election) may independently contribute to increased firearm injury and death during 2020.

## Conclusion

The effects of pandemic-related school closures on pediatric firearm encounters among children 5 to 17 are heterogeneous by race/ethnicity and age, reflecting underlying differences in household firearm ownership, safe storage practices, and neighborhood exposures to gun violence. There is a critical need to improve safe firearm storage in households with children and advance policies that address the root causes of gun violence in Black communities.

## Data Availability

Data contain personal health information from hospital records and are not publicly available.

## Acknowledgements

We thank Kirsty Clark for her review of the manuscript and attendees of the Vanderbilt University Medical Center Pediatrics Research Conference and Health Policy Works in Progress forums for their critical feedback on earlier stages of this project. This work was supported in part by the William Long Fellowship to Dr. Gastineau.

