## Appendix for "Trends in Pediatric Firearm-Related Encounters during the COVID-19 Pandemic by Age Group, Race/Ethnicity, and Schooling Mode in Tennessee"

| Appendix Table 1. Robustness Check 1: Incident Rate Ratios for Schooling Mode on Monthly Pediatric Firearm-Related Encounters with One Month Backward Shift in Timing of School Mode Changes | | | | | | | | |
| --- | --- | --- | --- | --- | --- | --- | --- | --- |
|  | Monthly Pediatric Firearm-Related Encounters | | | | | | | |
|  |  |  | By Race/Ethnicity | |  | By Age Group | | |
|  | Overall^a^ |  | Non-Hispanic Black | Non-Hispanic White |  | Age 0-11 | Age 12-15 | Age 16-17 |
| Schooling Mode |  |  |  |  |  |  |  |  |
| Pre-Pandemic (1/2018-2/2020) [reference] | -- |  | -- | -- |  | -- | -- | -- |
| Closed (3/2020-7/2020) | 1.333*** |  | 1.595*** | 1.277* |  | 2.242*** | 1.463*** | 0.966 |
|  | (0.092) |  | (0.148) | (0.130) |  | (0.360) | (0.151) | (0.0993) |
| Virtual/Hybrid (8/2020-1/2021) | 1.350* |  | 1.472** | 1.578+ |  | 1.035 | 1.898*** | 1.074 |
|  | (0.181) |  | (0.203) | (0.423) |  | (0.575) | (0.313) | (0.151) |
| In-Person (2/2021-12/2021) | 1.322** |  | 1.841*** | 0.512*** |  | 1.606* | 1.115 | 1.210* |
|  | (0.0847) |  | (0.182) | (0.0902) |  | (0.307) | (0.104) | (0.102) |
| Months (N) | 48 |  | 48 | 48 |  | 48 | 48 | 48 |
| Month Fixed Effects | Yes |  | Yes | Yes |  | Yes | Yes | Yes |
| Note: + p<0.1, * p<0.05, ** p<0.01, *** p<.001. Incident rate ratios are exponentiated coefficients. Robust standard errors in parentheses. ^a^ All models are predicted using a standard Poisson model with month of year fixed effects and robust standard errors. | | | | | | | | |

| Appendix Table 2. Robustness Check 2: Incident Rate Ratios for Schooling Mode on Monthly Pediatric Firearm-Related Encounters with One Month Forward Shift in Timing of School Mode Changes | | | | | | | | | | | | | | | | |
| --- | --- | --- | --- | --- | --- | --- | --- | --- | --- | --- | --- | --- | --- | --- | --- | --- |
|  | | Monthly Pediatric Firearm-Related Encounters | | | | | | | | | | | | | | |
|  | |  | |  | | By Race/Ethnicity | | | |  | | By Age Group | | | | |
|  | | Overall^a^ | |  | | Non-Hispanic Black | | Non-Hispanic White | |  | | Age 0-11 | | Age 12-15 | | Age 16-17 |
| Schooling Mode | |  | |  | |  | |  | |  | |  | |  | |  |
| Pre-Pandemic (1/2018-2/2020) [reference] | | -- | |  | | -- | | -- | |  | | -- | | -- | | -- |
| Closed (3/2020-9/2020) | | 1.428*** | |  | | 1.594*** | | 1.521*** | |  | | 2.502*** | | 1.796*** | | 0.941 |
|  | | (0.0920) | |  | | (0.138) | | (0.159) | |  | | (0.439) | | (0.193) | | (0.0875) |
| Virtual/Hybrid (10/2020-3/2021) | | 1.137 | |  | | 1.615** | | 0.915 | |  | | 0.529 | | 1.378+ | | 1.127 |
|  | | (0.171) | |  | | (0.267) | | (0.365) | |  | | (0.283) | | (0.253) | | (0.172) |
| In-Person (4/2021-12/2021) | | 1.289*** | |  | | 1.800*** | | 0.577*** | |  | | 1.863*** | | 1.137 | | 1.254** |
|  | | (0.0733) | |  | | (0.181) | | (0.0955) | |  | | (0.317) | | (0.102) | | (0.102) |
| Months (N) | | 48 | |  | | 48 | | 48 | |  | | 48 | | 48 | | 48 |
| Month Fixed Effects | | Yes | |  | | Yes | | Yes | |  | | Yes | | Yes | | Yes |
| Note: + p<0.1, * p<0.05, ** p<0.01, *** p<.001. Incident rate ratios are exponentiated coefficients. Robust standard errors in parentheses. ^a^ All models are predicted using a standard Poisson model with month of year fixed effects and robust standard errors. | | | | | | | | | | | | | | | | |
| Appendix Table 3. Robustness Check 3: Incident Rate Ratios for Schooling Mode on Monthly Pediatric Firearm-Related Encounters with Exclusion of March 2020 | | | | | | | | | | | | | | | | |
|  | Monthly Pediatric Firearm-Related Encounters | | | | | | | | | | | | | | | |
|  |  | |  | | By Race/Ethnicity | | | |  | | By Age Group | | | | | |
|  | Overall^a^ | |  | | Non-Hispanic Black | | Non-Hispanic White | |  | | Age 0-11 | | Age 12-15 | | Age 16-17 | |
| Schooling Mode |  | |  | |  | |  | |  | |  | |  | |  | |
| Pre-Pandemic (1/2018-2/2020) [reference] | -- | |  | | -- | | -- | |  | | -- | | -- | | -- | |
| Closed (4/2020-8/2020) | 1.466*** | |  | | 1.569*** | | 1.453** | |  | | 2.735*** | | 1.694*** | | 1.011 | |
|  | (0.112) | |  | | (0.133) | | (0.178) | |  | | (0.471) | | (0.244) | | (0.0865) | |
| Virtual/Hybrid (9/2020-1/2021) | 1.135 | |  | | 1.489** | | 1.105 | |  | | 0.319*** | | 1.546* | | 1.045 | |
|  | (0.165) | |  | | (0.223) | | (0.381) | |  | | (0.111) | | (0.283) | | (0.162) | |
| In-Person (2/2021-12/2021) | 1.295*** | |  | | 1.855*** | | 0.552*** | |  | | 1.951*** | | 1.138 | | 1.258** | |
|  | (0.0718) | |  | | (0.186) | | (0.0886) | |  | | (0.334) | | (0.0998) | | (0.0994) | |
| Months (N) | 47 | |  | | 47 | | 47 | |  | | 47 | | 47 | | 47 | |
| Month Fixed Effects | Yes | |  | | Yes | | Yes | |  | | Yes | | Yes | | Yes | |
| Note: + p<0.1, * p<0.05, ** p<0.01, *** p<.001. Incident rate ratios are exponentiated coefficients. Robust standard errors in parentheses. ^a^ All models are predicted using a Poisson model with month of year fixed effects and robust standard errors. | | | | | | | | | | | | | | | | |

| Appendix Table 4. Robustness Check 4: Incident Rate Ratios for Schooling Mode on Monthly Pediatric Firearm-Related Encounters with Exclusion of January 2018 Kentucky High School Shooting Victims | | | | | | | | |
| --- | --- | --- | --- | --- | --- | --- | --- | --- |
|  | Monthly Pediatric Firearm-Related Encounters | | | | | | | |
|  |  |  | By Race/Ethnicity | |  | By Age Group | | |
|  | Overall^a^ |  | Non-Hispanic Black | Non-Hispanic White |  | Age 0-11 | Age 12-15 | Age 16-17 |
| Schooling Mode |  |  |  |  |  |  |  |  |
| Pre-Pandemic (1/2018-2/2020) [reference] | -- |  | -- | -- |  | -- | -- | -- |
| Closed (3/2020-8/2020) | 1.459*** |  | 1.626*** | 1.473*** |  | 2.756*** | 1.693*** | 0.981 |
|  | (0.0995) |  | (0.138) | (0.167) |  | (0.451) | (0.210) | (0.0859) |
| Virtual/Hybrid (9/2020-1/2021) | 1.179 |  | 1.489** | 1.232 |  | 0.319*** | 1.546* | 1.128 |
|  | (0.149) |  | (0.223) | (0.362) |  | (0.111) | (0.283) | (0.146) |
| In-Person (2/2021-12/2021) | 1.302*** |  | 1.855*** | 0.561*** |  | 1.951*** | 1.138 | 1.271** |
|  | (0.0705) |  | (0.184) | (0.0878) |  | (0.334) | (0.0997) | (0.100) |
| Months (N) | 48 |  | 48 | 48 |  | 48 | 48 | 48 |
| Month Fixed Effects | Yes |  | Yes | Yes |  | Yes | Yes | Yes |
| Note: + p<0.1, * p<0.05, ** p<0.01, *** p<.001. Incident rate ratios are exponentiated coefficients. Robust standard errors in parentheses. ^a^ All models are predicted using a standard Poisson model with month of year fixed effects and robust standard errors. | | | | | | | | |
